# Using rapid (point-of-care) tests for COVID-19: A decision analysis comparing the expected benefit of two screening strategies

**DOI:** 10.1101/2021.04.15.21255569

**Authors:** Raymond H. Baillargeon, Xavier Seyer, Éricka Bernard-Bédard

## Abstract

**Background:** Rapid tests for COVID-19 could be used to augment the otherwise limited laboratory-based testing capacity, but there are concerns that their utility may be compromised by their limited accuracy. The objective of this article is to compare the expected benefit (EB) of two screening strategies, one with rapid tests (SwRT) and another one without rapid tests 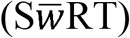.

**Methods:** We performed a decision analysis, with the overall EB defined as the proportion of correctly identified individuals minus the proportion of incorrectly identified individuals. Accordingly, the SwRT strategy will be deemed a better screening strategy if its lesser EB for COVID-19 free individuals is more than compensated by its greater EB for COVID-19 individuals. Otherwise, it will not.

**Results:** As expected, the EB for COVID-19 individuals was greater for the SwRT strategy, with a far superior ability to rule out the presence of COVID-19. In fact, under the scenario of interest (i.e., 8000 ID Now rapid tests in addition to 28185 lab-based RT-PCR tests), it identified almost 16% more COVID-19 individuals than the 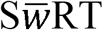 strategy. In addition, the EB for COVID-19 free individuals was the same for both strategies, with a perfect ability at ruling in the presence of COVID-19.

**Conclusion:** The SwRT strategy identified more COVID-19 individuals and this gain was not obtained at the detriment of COVID-19 free individuals who were equally well identified by both strategies. Hence, the SwRT strategy is a better screening strategy for COVID-19. It represents an opportunity to curtail the spread of SARS-CoV-2 that we may not afford to miss with new more contagious variants becoming more and more common in Canada.

---

The timely identification of COVID-19 individuals via testing is key to limit the spread of SRAS-CoV-2 in the community.^1^ This may be especially true with new more contagious variants becoming more common in Canada, and the possibility of a ‘third wave’ looming at the horizon. Expert committees at the federal level^2^ and in at least two provinces (i.e., Québec, Alberta)^3,4^ have considered the possibility of using rapid point-of-care (molecular and antigen) tests for COVID-19. But so far, efforts to implement rapid tests have fallen short as very few of the rapid tests being distributed (freely) to the provincial/territorial jurisdictions are being used. For example, in Québec, Le Devoir was recently reporting than less than 2% are.^5^

Rapid tests could be used to augment the otherwise limited lab-based testing capacity, but there are concerns that their utility may be compromised by their limited accuracy. If the goal is to have the most efficient screening strategy (see expected benefit benchmark below) then the question becomes whether the potential benefits of using rapid tests outweigh the costs? The objective of this article is to conduct a decision analysis to compare the overall expected benefit [EB; proportion of correctly identified individuals - proportion of incorrectly identified individuals] of two COVID-19 screening strategies, one with rapid tests (SwRT) and another one without rapid tests 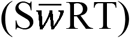. On the one hand, we predict the EB for COVID-19 individuals (i.e., proportion of true positive outcomes - proportion of false negative outcomes/not tested) to be higher for the SwRT strategy. On the other hand, we predict the EB for COVID-19 free individuals (i.e., proportion of true negative outcomes/not tested - proportion of false positive outcomes) to be smaller for the SwRT strategy. The SwRT strategy will be deemed a better screening strategy if its lesser EB for COVID-19 free individuals is more than compensated by its greater EB for COVID-19 individuals. Otherwise, it will not.

## Methods

One way to compare the EB of the two screening strategies is to use decision analysis.^6^ This formal process utilizes estimates from empirical studies to determine what the best strategy is. This information might include a test’s sensitivity (SE) and specificity (SP) estimates obtained from a diagnostic test accuracy (DTA) study. Each strategy is depicted as a probability tree (diagram) that includes all possible events (e.g., obtaining a positive rapid test result for COVID-19 individuals). Events constitute the tree’s branches. Mutually exclusive and exhaustive events appear at the right of chances nodes (represented by circles) together with their corresponding probabilities. Also, a probability tree includes all possible outcomes (e.g., being correctly identified as having COVID-19--true positive outcome). They appear at the far right of the probability tree. There is one for each possible sequence of events (i.e., paths). The (path) probability of a particular outcome is obtained by multiplying the probabilities associated with all the events leading to the outcome in question. (The probability of a particular event being conditional on all the events the precede it in the sequence.) Finally, the two strategies are linked by a decision node (represented by a square) and together they constitute a decision tree.

Empirical estimates such as a test’s sensitivity (SE) and specificity (SP) often need to be calibrated (on a scale with values ranging from 0 to 1) before they can be used to assign probabilities to events in a probability tree.^7^ An estimate of a test’s SE depends on the proportion of individuals who obtained a positive test result in that particular study (i.e., the latter corresponding to the value of SE expected by chance alone). Also, an estimate of a test’s SP depends on the proportion of individuals who obtained a negative test result in that particular study (i.e., the latter corresponding to the value of SP expected by chance alone). Following Kraemer,^8^ SE and SP can be calibrated using the weighted kappa coefficients k(1,0) and k(0,0), respectively. The coefficient k(1,0) is equal to (SE - SE expected by chance alone) / (1 - SE expected by chance alone). Similarly, k(0,0) is equal to (SP - SP expected by chance alone) / (1 - SP expected by chance alone). These calibrated values can then be used to obtain probabilities of events (e.g., obtaining a positive rapid test result for COVID-19 individuals) whatever the proportion of individuals with a positive/negative test result might be under the scenario of interest.

## Results

Table 1 presents the information required to estimate the EB of the two screening strategies.

**Table 1.** Information used to estimate the expected benefit of the two screening strategies

| Information | Estimate |  |
| --- | --- | --- |
| Number of RT-PCR tests used (per day) | 28185 |  |
| Number of ID Now rapid tests used (per day) | 8000 |  |
| Proportion of RT-PCR tests | 0.7789 |  |
| Relative prevalence of COVID-19 | 0.0361 |  |
| Accuracy of the RT-PCR tests | Perfect |  |
| Sensitivity of the ID Now, calibrated value [i.e., $k(1,0)$] | 0.5469 | |
| Specificity of the ID Now, calibrated value [i.e., $k(0,0)$] | 1.0000 | |
| Odds ratio for the marginal probabilities (i.e., odds of COVID-19 / odds of a positive ID Now test result) | 1.8283 |  |
| Proportion of positive ID Now test results | 0.0201 |  |
| Sensitivity of the ID Now, uncalibrated value | 0.5560 |  |
| Specificity of the ID Now, uncalibrated value | 1.0000 |  |

First, under the scenario of interest, the proportion of lab-based reverse transcription polymerase chain reaction (RT-PCR) tests was estimated at .779. For the reference period going from February 1^st^ to February 10^th^, 281848 lab-based RT-PCR tests were performed at screening centers in Québec (i.e., an average of approximately 28185 tests per day) according to the Institut National de Santé Publique du Québec (INSPQ).^9^ Also, from a recent press release from the Ministère de la Santé et des services sociaux (Québec’s Health Ministry),^10^ we postulated that, under the SwRT strategy, an additional 8000 Abbott ID Now tests could be performed on a daily basis. The ID Now is an isothermal nucleic acid amplification test that has been approved for commercial/diagnostic use by Health Canada (30 September 2020).^11^ It is one of many rapid tests for COVID-19 that are being distributed (at no cost) by the Public Health Agency of Canada to the provincial/territorial jurisdictions.

Second, the relative prevalence of COVID-19 (at screening centers) was estimated at .0361 using data from the INSPQ.^9^ In fact, for the mentioned above reference period, there were 10183 confirmed cases and 271665 non confirmed cases.

Third, the accuracy of lab-based RT-PCR tests for COVID-19 was assumed to be perfect.

Fourth, the calibrated values of the ID Now’s SE and SP were estimated from a DTA study conducted by Moore and his colleagues.^12^ According to a recent Cochrane DTA systematic review,^13^ this is the only DTA study of the ID Now that used two lab-based RT-PCR tests. It is generally accepted that, to be considered valid for DTA purpose, a negative test result for COVID-19 needs to be confirmed by two lab-based RT-PCR tests.^13^ We re-analyzed Moore et al.’s data for patients with conclusive/valid test results on all three tests (n = 196 / 200). By applying the above rule, we obtained the following results (i.e., number of): true positives = 94; false negatives = 29; false positives = 0; and true negatives = 73. The calibrated value of SE was estimated at .5469 [i.e., (0.7642 - 0.4796) / (1.0 - 0.4796)], where 0.7642 is the uncalibrated value of SE and 0.4796 is the value of SE expected by chance alone (i.e., the proportion of patients who obtained a positive ID Now test result). Also, the calibrated value of SP was estimated at 1.0 [i.e., (1.0 - 0.5204) / (1 - 0.5204)], where 1.0 is the uncalibrated value of SP and 0.5204 is the value of SP expected by chance alone (i.e., the proportion of patients who obtained a negative ID Now test result). Finally, the odds ratio for the marginal probabilities was estimated at 1.8283 [i.e., (1.6849 / .9216)], where 1.6849 (i.e., 123 / 73) are the odds of COVID-19 based on the two RT-PCR tests and .9216 (i.e., 94 / 102) are the odds of a positive ID Now test result.

Fifth, the corresponding proportion of individuals with a positive ID Now test result (i.e., .0201) was determined by solving the equation: [(.0361 / .9639) / odds of a positive ID Now test result = 1.8283], where .0361 is the relative prevalence of COVID-19 and 1.8283 is the odds ratio for the marginal probabilities (see above). The calculated odds of a positive ID Now test result (i.e., .0205) was then transformed into the probability of a positive ID Now test using the standard formula [i.e., odds / (1 + odds)]. The corresponding (uncalibrated) value of SE (i.e., .5560) was obtained by solving the equation: [(SE - .0201) / (1 - .0201) = .5469], where .0201 is the proportion of individuals with a positive ID Now test result and .5469 is the calibrated value of SE. Also, the corresponding (uncalibrated) value of SP (i.e., 1.0) was equal to its calibrated value of 1.0.

Figure 1 (upper part) depicts the SwRT strategy. For the lab-based RT-PCR tests there are only two different sequence of events and their corresponding appropriate outcomes (i.e., true positive and true negative). For the rapid ID Now tests there are four different sequences of events and their corresponding appropriate/not appropriate outcomes (i.e., true positive, false negative, false positive and true negative). At the far right of the diagram appear the probabilities associated with the six outcomes in question. (These path probabilities sum to one.) Also, Figure 1 (lower part) depicts the 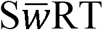 strategy. Not using the ID Now amounts to giving a negative test result to all individuals whether they have COVID-19 or not (i.e., two sequence of events). For that reason, the two corresponding outcomes are labelled ‘false negative’ (not appropriate) and ‘true positive’ (appropriate), respectively.

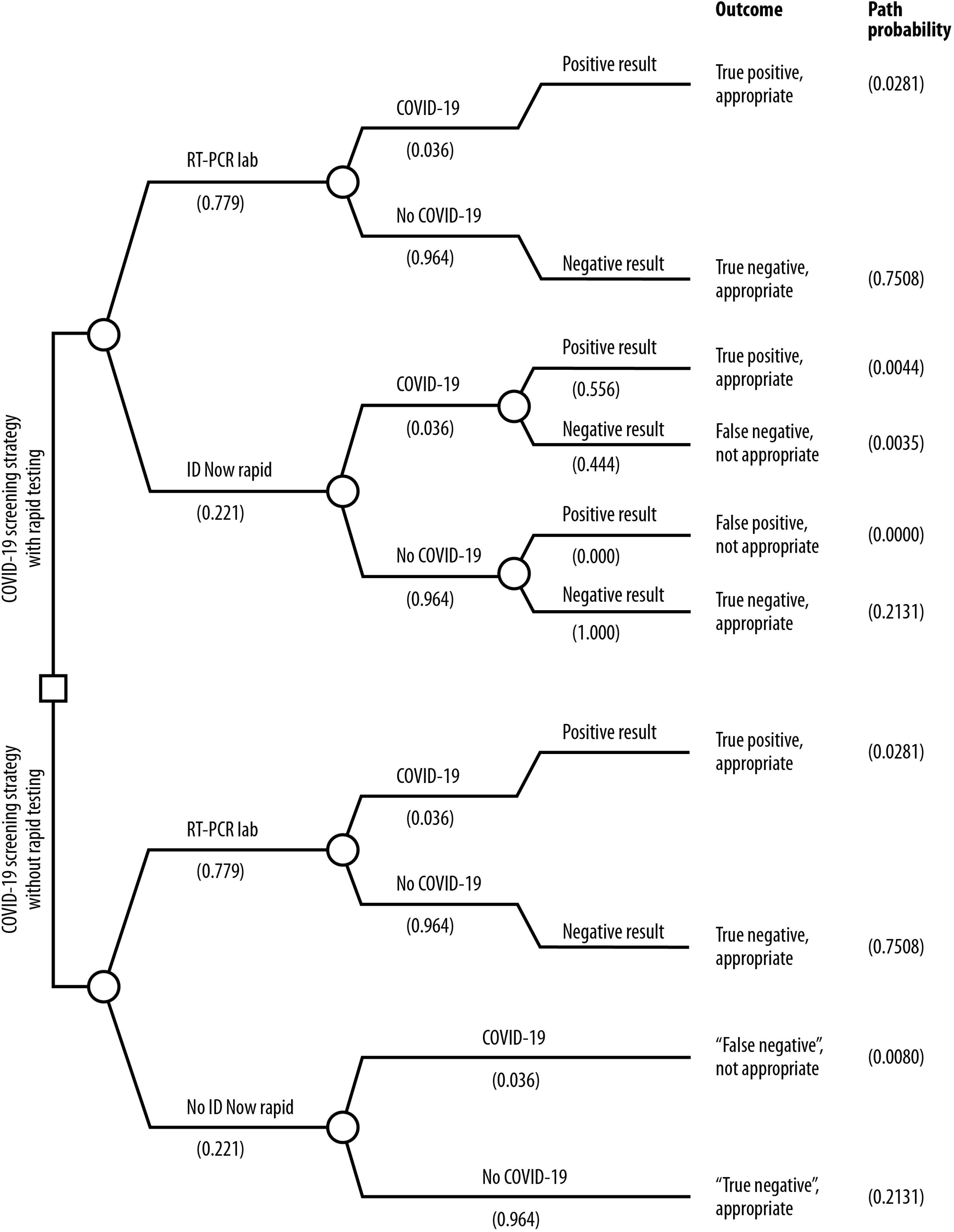

Table 2 presents, for each screening strategy, the averaged-out probabilities for the four outcomes of interest. First, as expected, the EB for COVID-19 individuals was greater for the SwRT (i.e., .0326 - .0035 = .029) than for the 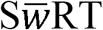 strategy (i.e., .0281 - .0080 = .020). In fact, the SwRT strategy’ ability to rule out the presence of COVID-19 was far superior. Under the SwRT strategy, 90.2% of COVID-19 individuals were correctly identified. In comparison, only 77.9% of COVID-19 individuals were correctly identified under the 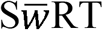. In other words, the SwRT strategy identified almost 16% more COVID-19 individuals. Under the scenario of interest where a total of 36185 tests per day would be administered (i.e., 28185 lab-based RT-PCR tests and 8000 ID Now tests) this represents a gain of approximately 161 confirmed cases per day (i.e., 1179 vs. 1018). Second, the EB for COVID-19 free individuals was the same for both strategies (i.e., .9639 - .0000 = .964), with a perfect ability at ruling in the presence of COVID-19. (Under the scenario of interest, the ID Now was considered a perfectly specific test.) Overall, the EB benefit was greater for the SwRT (i.e., .029 + .964 = .993) than for the 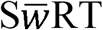 strategy (i.e., .020 + .964 = .984). The former identified more COVID-19 individuals and this gain was not obtained at the detriment of COVID-19 free individuals who were equally well identified by both strategies.

**Table 2.** Averaged out probabilities for the four outcomes of interest under the two screening strategies

|  |  |  |  |  |
| --- | --- | --- | --- | --- |
|  | Screening strategy with rapid tests (SwRT) |  |  |  |
|  | Test result |  |  |  |
|  | Positive | Negative | Total |  |
| Disease status |  |  |  |  |
| COVID-19 | .0326 | .0035 | .0361 |  |
| No COVID-19 | .0000 | .9639 | .9639 |  |
| Total | .0326 | .9674 | 1.000 |  |
| | Screening strategy without rapid tests (S$\bar{w}$RT) | | | |
|  | Test result |  |  |  |
|  | Positive | Negative/Not tested | Total |  |
| Disease status |  |  |  |  |
| COVID-19 | .0281 | .0080 | .0361 |  |
| No COVID-19 | .0000 | .9639 | .9639 |  |
| Total | .0281 | .9719 | 1.000 |  |

Table 3 compares the EB of the two screening strategies under a broad range of values for the (unique) probabilities assigned to the different events that appear in the decision tree (see Figure 1). The evidence of favor of the SwRT strategy remains basically the same, except for a change in the number of rapid tests being administered. In fact, the difference between the two strategies in the proportion of COVID-19 individuals being correctly identified was estimated at only 7.89% if fewer (i.e., 4000 instead of 8000) rapid tests were administered--a gain of approximately 80 confirmed cases per day. In contrast, this differential was estimated at 23.67% if more (i.e., 12000 instead of 8000) rapid tests were administered--a gain of approximately 241 confirmed cases per day.

**Table 3.** Sensitivity analysis for the proportion of correctly identified COVID-19 individuals under the two screening strategies

|  |  |  |
| --- | --- | --- |
|  | Relative prevalence of COVID-19 (.036; se = .002; cv = 5.12%) |  |
| | Lower bound of 95% large-sample CI (0.033)<br>SwRT/S$\bar{w}$RT | Upper bound of 95% large-sample CI (0.040)<br>SwRT/S$\bar{w}$RT |
|  | .9016/.7789 | .9021/.7789 |
|  | Proportion of lab-based RT-PCR tests (.779 or 8000 ID Now tests) |  |
| | (.876 or 4000 ID Now tests)<br>SwRT/S$\bar{w}$RT | (.701 or 12000 ID Now tests)<br>SwRT/S$\bar{w}$RT |
|  | .9448/.8757 | .8674/.7014 |
|  | Odds ratio for the marginal probabilities (Jackknife estimate = 1.828; se = .014; cv = .75%) |  |
| | Lower bound of 95% Jackknife CI (1.802)<br>SwRT/S$\bar{w}$RT | Upper bound of 95% Jackknife CI (1.855)<br>SwRT/S$\bar{w}$RT |
|  | .9036/.7789 | .9001/.7789 |
|  | Calibration of SE [i.e., k(1,0)] (Jackknife estimate = .547; se = .004; cv = .76%) |  |
| | Lower bound of 95% Jackknife CI (.539)<br>SwRT/S$\bar{w}$RT | Upper bound of 95% Jackknife CI (.555)<br>SwRT/S$\bar{w}$RT |
|  | .9001/.7789 | .9037/.7789 |

Note. Under the scenario of interest, the proportion of correctly identified COVID-19 individuals was estimated at .9019 and .7789 under the SwRT and S $\bar{w}$ RT strategy, respectively. Because $k(0,0) / k(1,0)$ = odds ratio for the marginal probabilities, the sensitivity results for SE and the marginal odds are mirror images of each other.
SwRT: screening with rapid tests.
S $\bar{w}$ RT: screening without rapid tests.

## Interpretation

Beyond practical considerations, there are still many unanswered questions surrounding the use of readily available rapid (point-of-care) tests to augment the otherwise limited laboratory-based testing capacity for COVID-19. The key question is whether the potential benefits of using rapid tests outweigh the costs. The goal of this article was to compare the EB of two screening strategies for COVID-19, one with rapid tests (SwRT) and another one without rapid tests 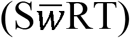. The results show that the ability to rule out the presence of COVID-19 was greater under the SwRT, with 16% more COVID-19 individuals being correctly identified--approximately 161 confirmed cases per day for a province like Québec with a target of 35,000 tests per day.^14^ Because the ability to rule in the presence of COVID-19 was the same for both strategies (i.e., 100%), this gain was not obtained at the detriment of COVID-19 free individuals. Hence, on all accounts, the SwRT strategy is a better screening strategy for COVID-19.

### How to optimize the effectiveness of a testing strategy that relies on rapid tests

Of course, if more rapid tests were administered, but also if a different, more sensitive, test was used instead of the ID Now. For instance, the rapid Cepheid Xpert Xpress molecular test has demonstrated a better clinical sensitivity in DTA studies.^13,15^ Also, it provides not only a qualitative result, but also a reading of the viral load of the patient (i.e., cycle threshold). This could greatly facilitate how we deal with negative rapid test results. For individuals with a low viral load (e.g., cycle threshold values above 30),^16^ a negative test result could be systematically investigated using a lab-based RT-PCR test. But confirming or not confirming a negative test result may not mater much in terms of limiting the outbreak because contagiousness/transmission is most likely to occur when the viral load peaks.^17^ In fact, in a recent simulation study, Larremore and his colleagues^18^ have demonstrated that (under some circumstances) rapid tests may be as good as lab-based tests when it comes to stop the transmission of SARS-CoV-2. At any event, individuals with a negative rapid test result should continue to adhere to the usual public health precautions (i.e., washing hands frequently, respecting social distancing, wearing masks and, if sick, staying at home and self isolate).

## Limitations

First, the accuracy of lab-based RT-PCR tests for COVID-19 was assumed to be perfect despite recent evidence that they may generate false-negative test results.^19-21^ Had this been taken into account, the differential between the two strategies would only have been greater. Second, the above-cited Cochrane DTA systematic review^13^ has since been updated,^22^ but the more recent DTA studies of the ID Now do not seem to have used, as recommended,^13^ two lab-based RT-PCR tests to confirm a negative test result. Hence, as far as we know, our reanalysis of the data from Moore et al.^12^ represents the best attempt to date to obtain calibrated values of the ID Now’s SE and SP. Third, this is a preprint version of this article, and therefore it has not been peer reviewed.

## Conclusion

Experts who have cautioned against using rapid tests have done so based on their lack of perfect accuracy. Paradoxically, not using rapid tests amounts to give a negative test results to all these individuals (including those with COVID-19) who could have been tested otherwise (had rapid tests been implemented). Not testing might be a way to avoid false-positive test results, but it is not an issue here, with the specificity of rapid tests being comparable to the one of lab-based RT-PCR tests. Thus, a screening strategy using (valid) rapid tests to augment our testing capacity can allow us to identify more COVID-19 individuals. Moreover, it does not do so at the detriment of COVID-19 free individuals. It is a net gain. It represents an opportunity that we may not afford to miss at a time where new more contagious variants are becoming more common in Canada. What are we waiting for?

## Data Availability

All the relevant data are available in the manuscript.

## Authors’ contributions

RHB conceived the study. All of the authors were involved in the data analysis. RHB wrote the first draft of the article and all the authors contributed to its critical revision. All of the authors approved the final version of the article and are accountable for all of its aspects.

## Acknowledgements

We would like to thank Nigèle Langlois for her help with searching the literature. Also, we would like to thank Marc Bélanger for his help with the figure. Finally, we would like to thank all with whom we exchanged correspondence while writing this paper.

